# Fecal iron quantification in a randomized controlled trial of *Lactiplantibacillus plantarum* ATCC 202195 in newborns in Dhaka, Bangladesh

**DOI:** 10.1101/2025.09.25.25336651

**Authors:** Afreen Z. Khan, Anjan K. Roy, Huma Qamar, Lisa G. Pell, Karen M. O’Callaghan, Shafiqul A. Sarker, Abdullah A. Mahmud, Rashidul Haque, Sharmin Akter, Shamima Sultana, Daniel E. Roth, Rubhana Raqib

## Abstract

**Background:** Probiotics may enhance host iron bioavailability, offering a strategy to address iron deficiency. Fecal iron may be a useful non-invasive biomarker of such effects in infants.

**Objective:** To examine the use of fecal iron quantification in a randomized placebo-controlled trial (RCT) of neonatal administration of *Lactiplantibacillus plantarum* ATCC 202195 (LP202195), with or without fructooligosaccharide (FOS), in Dhaka, Bangladesh.

**Methods:** Fecal iron quantification using atomic absorption spectrometry (AAS) was optimized using standards and reference materials, and pilot-tested using pooled stool aliquots (n=32) from an observational cohort of young infants in Bangladesh (aged 0-64 days). The optimized AAS assay was then applied to individual stool samples collected at 14 days of age (n=307) in a RCT in which newborns aged 0-4 days were randomly allocated to one of five groups: placebo, 1-or 7-day regimens of LP202195, with or without FOS. Serum ferritin was measured at 2 months postnatal age (n=251). Effects of the 1-and 7-day LP202195 regimens were estimated using linear regression and expressed as mean percent differences relative to placebo, with 95% confidence intervals (95%CI).

**Results:** The optimized AAS fecal iron assay had acceptable accuracy (91-99%), precision (within-and between-run coefficients of variation <10%), and recovery (93-112%), with a reportable range of 0.2 to 80 mg Fe per 100 g dry stool. In pooled samples from the observational cohort, fecal iron varied with age and feeding status. In the RCT, fecal iron concentrations did not significantly differ following1-day (% difference=9.8%, 95%CI:-19%, 49%; P=0.54) or 7-days (% difference=-6.1%, 95%CI:-31%, 28%; P=0.69) of LP202195 administration, versus placebo (geometric mean concentration=4.3mg Fe/100g dry stool (95%CI:3.3, 5.6); n=53). Inferences were unchanged when groups were disaggregated by FOS co-administration (P>0.05 for all). Similarly, there were no effects of LP202195 on serum ferritin at 2 months of age (P>0.05 for all).

**Conclusions:** Fecal iron quantification by AAS was valid and feasibly implemented in a trial of neonatal administration of *Lactiplantibacillus plantarum* ATCC 202195. However, the assay is resource-intensive and may not be more informative than conventional measures of iron status when studying the effects of probiotics/synbiotics on iron bioavailability.

**Clinical Trial Registry:** ClinicalTrials.gov identifier: NCT05180201

## Introduction

Iron deficiency (ID) is common during early childhood and can have irreversible adverse effects on neurodevelopment[1, 2]. Daily oral iron supplementation reduces the risk of ID, anemia, and iron deficiency anemia in children aged 4 to 23 months[3]. However, iron stores in term, exclusively breastfed infants are generally assumed to be sufficient until 4 to 6 months of age[4–6]; for this reason, the World Health Organization (WHO) only recommends routine iron supplementation for infants aged 6 months and older in low-and middle-income countries (LMICs) where there is a high prevalence of ID and anemia[7]. WHO also recommends a daily intake of 30-60mg elemental iron along with 0.4mg folic acid in pregnancy, starting soon after conception[8]. Yet, many infants in LMICs may be iron deficient at an earlier age (< 6 months) due to the greater risk of preterm birth and low birthweight, which are associated with lower iron stores at birth[9]. Additionally, iron absorption is more likely to be impaired in populations living in LMICs due to a higher prevalence of infection and chronic inflammation[10, 11], thereby contributing to the risk of ID. Since most intestinal bacteria rely on iron for survival and virulence [12], there is competition between intestinal microbes and their human host for iron in the gastrointestinal tract, whereby excess intraluminal iron may promote the proliferation of pathogenic bacteria rather than benefit the host[13].

While iron supplementation and fortification strategies have shown benefits in reducing the prevalence of ID and anemia in infants and young children[3, 14], the safety of iron-containing interventions has been questioned in some LMIC settings with high rates of malaria and/or other infections, where clinical trials have linked exogenous iron to an increased risk of serious adverse events including deaths and hospital admissions[15], and bloody diarrhea[16]. Iron interventions may negatively impact the gut microbiota composition by increasing the abundance of potentially pathogenic bacteria and decreasing the abundance of commensal bacteria, which may contribute to intestinal inflammation[13, 17]. Alternative interventions that safely improve iron status are therefore of interest and may be particularly applicable in early infancy before the age when routine iron supplementation is recommended. Lactobacilli probiotics are iron-independent and have the potential to improve iron absorption in the human intestine[18–20] through multiple pathways including alteration of gut microbiota composition[19], increases in lactic acid and short-chain fatty acids that may lower intestinal pH[21, 22], and the production of other metabolites that promote iron solubility[23]. The co-administration of iron and lactobacilli-containing probiotic regimens has been shown to improve iron absorption in animal models[24] and in human females of reproductive age[18, 25, 26], but such benefits have not been observed to date in pediatric populations[27, 28]. Little is known about the effect of probiotic administration alone on gastrointestinal iron absorption without co-administration of an exogenous iron source, and no studies have explored the effect of probiotic administration in early infancy on iron status or markers of iron absorption or excretion.

Fecal iron concentration is a non-invasive measure of ingested unabsorbed iron, but is not a direct marker of iron status or iron absorption. However, fecal iron may be valuable in studies of microbiome-iron interactions because the same stool samples collected for microbiome analyses may be used for iron quantification, potentially alleviating the need for blood collection or use of direct yet more expensive methods such as stable isotope administration[29]. While several techniques for fecal iron quantification have been described in the literature, none are routinely used in the context of microbiome or probiotic research[29]. In the present study, we first optimized and validated an atomic absorption spectrometry (AAS) method for fecal iron quantification in stool samples from a cohort of infants in Bangladesh. Then, in a sub-study of a randomized placebo-controlled trial (RCT) of *Lactiplantibacillus plantarum* ATCC 202195 (LP202195)[30], we applied this method to examine if LP202195 with and/or without fructooligosaccharide (FOS), for 1-or 7-days, affects stool iron content at 14 days postnatal age and serum ferritin at 60-days postnatal age in young infants in Dhaka, Bangladesh. In the context of the RCT, whereby average prenatal endowment of iron stores and postnatal iron intake were assumed to be similar across the intervention groups, we expected that a relatively lower stool iron concentration in infants in the intervention group compared to placebo would indicate reduced intraluminal excess iron and lower stool iron losses, thereby serving as an indirect indicator of greater fractional iron absorption. We further anticipated that an increase in iron absorption following probiotic intervention would manifest as a greater mean ferritin concentration in the intervention group relative to placebo at 2 months of age.

## Methods

### Part A: Fecal iron assay optimization and validation

Stool samples for assay optimization were randomly selected from infants enrolled in the SEPSiS observational cohort study (ClinicalTrials.gov ID: NCT04012190) for which recruitment, enrollment and stool collection procedures have been previously described (Section A in Supplementary Material)[31]. Samples collected from 0 to 60 days of age (n=130) were pooled and divided into 1-2 g aliquots. Reference materials and spiked samples were also used in optimization procedures (Table S1 in Supplementary Material).

The sample processing and AAS method was adapted from Pizarro *et al.*[32] and Fairweather-Tait *et al*[33]. Aliquots were placed in a freeze dryer for 3-4 days, following which the weight of the dry stool was determined. Specimens were transferred into re-usable crucibles which were washed thoroughly between uses using chromic acid followed by nitric acid. The crucibles were organized inside a muffle furnace and specimens ashed at increasing temperatures as follows: 100°C for 1 hour, 200°C for 1 hour, 500°C for 2 hours and 600°C for 2 hours. The ashing procedure was repeated every day for up to 3 days, and considered complete when the sample was entirely white (i.e., lacking black residue). Spiked stool samples, rice flour and turmeric powder were lyophilized and ashed in a similar manner. Each ashed sample was reconstituted in 2mL of 0.2N hydrochloric acid (HCl), then further diluted and aspirated into the flame of the ASC-7000 Atomic Absorption Autosampler (Shimadzu, Maryland, USA). Initial dilutions were 5-fold, but increased to 10-, 20-, or 40-fold until an absorbance value in the reportable range was obtained. Absorbances were read using a wavelength range of 248.3nm and iron concentrations were determined using a standard curve generated using five dilutions of a stock solution of known iron concentration, with final concentrations ranging from 0.4 to 4.0mg/L (Figure S1).

Exploratory analyses indicated that results were not acceptably reproduced when specimen weights were below 0.25g (dry weight of stool); also, it was not expected that dry weights greater than ∼0.3g would be readily available in future studies using individual infant stool aliquots. Therefore, we decided to perform all subsequent assays using at least 0.3g lyophilized stool.

The assay performance characteristics included lower limit of quantification (LLOQ), lower limit of detection (LOD), precision expressed as coefficients of variation (CV%), accuracy (percentage error from the known standard) and iron recovery (Section B in Supplementary Material).

A pilot study was conducted using an additional set of 158 stool samples from 123 infants (pooled into 32 samples) from the SEPSiS observational cohort. Samples were selected using a stratified approach with 4 variables (details in Supplementary Material, Section C):

i. Age (enrolment, and days 14 and 60 +/-7-days for each age time point)
ii. Infant feeding pattern (exclusive or predominant breastfeeding, partial breastfeeding, and no breastfeeding)
iii. Sex (Male vs. Female)
iv. Neonatal nutritional status (Yes or No for underweight)

Fecal iron quantification was performed as described above and expressed as mg of iron/100g dry weight of stool for each pool. Pools were combined for analysis based on shared characteristics (Supplementary Table S5), and iron concentration was expressed as mean (standard deviation) with 95% Confidence Intervals (95% CI).

Part B: Fecal iron quantification in a randomized controlled trial

### Study design and participants

This study was nested within a randomized, placebo-controlled trial of neonatal oral administration of LP202195 with or without FOS, for 1-or 7-days (ClinicalTrials.gov ID: NCT05180201), referred to below as the SEPSiS LP trial, for which recruitment, eligibility screening, and enrollment procedures have been previously described (Section A in Supplementary Material)[30]. In brief, infants were recruited, screened, and enrolled at two government hospitals in Dhaka, Bangladesh: Maternal and Child Health Training Institute (MCHTI) and Mohammadpur Fertility Services and Training Centre (MFSTC). Participants were randomly assigned to one of five intervention groups: placebo; LP202195 without FOS for 1 day (LP1); LP202195 with FOS for 1 day (LP1+FOS); LP202195 without FOS for 7 days (LP7); or LP202195 with FOS for 7 days (LP7+FOS). Infants were eligible to participate in the primary trial if they were: delivered at MCHTI or MFSTC; 0 to 4 days of age (day of birth was defined as day 0); weighed ≥1500g at birth; orally feeding at the time of eligibility assessment; and, their caregiver intended to reside within the study catchment area until the infant was ≥60 days postnatal age. To be eligible for inclusion in this nested sub-study, SEPSiS LP trial participants (infants) were required to have a stored stool sample aliquot collected at approximately 14 days postnatal age timepoint (although eligible samples were collected up to 4 weeks of age) and/or a stored blood sample aliquot collected as scheduled for the 60-day visit. The outcomes in this sub-study, stool iron and serum ferritin, were both specified as secondary outcomes in the SEPSiS LP trial protocol. Stool iron was quantified in samples collected from the first 307 infants enrolled in the SEPSiS LP trial for whom at least 2g of a day 14 stool sample was available for stool iron analysis.

### Intervention

Active investigational product (IP) vials consisted of either 10^9^ colony forming units of LP202195 with 150 mg FOS and 100 mg of maltodextrin (synbiotic regimen), or 10^9^ colony forming units of LP202195 and 250 mg of maltodextrin only (probiotic regimen). Placebo-containing IP vials contained 250 mg of maltodextrin. IPs were preferentially reconstituted in 3mL of human milk (hand-expressed from mother); alternatively, 3mL of sterile water or a mixture of water and human milk (total 3 mL) were also permitted. IP was orally administered to infants by study personnel and efforts were made to administer the IP once per day for seven consecutive days up to a maximum of seven doses per participant between 0-21 days of age[30].

### Data and specimen collection

All data were collected in the SEPSIS LP trial during hospital/clinic and home visits, via standard data collection forms and clinical measurements, using an electronic data capture system[30]. Study personnel used standard questionnaires at enrollment and/or hospital records to collect information related to maternal age at delivery, gestational age at delivery, mode of delivery, gravidity, birthweight, infant sex, maternal education, maternal occupation, enrollment site and asset index. Infant feeding status was defined using feeding-related data collected up to the day 14 study visit. Asset index of the participants was based on infants enrolled in both the SEPSiS LP trial (including infants in this sub-study) as well as the linked SEPSiS observational cohort. Infant stool samples were routinely collected at enrollment (postnatal days 0-4) and on postnatal days 4, 5 or 7 (randomly assigned), 10, 14, 21, 28, 42, 60, 90, and 180 (rescheduled when necessary)[30]. The 14-day postnatal age sample was selected for stool iron assessment as it was expected that most infants would have completed the probiotic/synbiotic or placebo intervention regimens prior to this age. All stool samples were collected by trained study personnel from a sanitized plastic sheet where they were homogenized, aliquoted, placed in the vapour phase of liquid nitrogen within 20 minutes of defecation, and transported to the International Centre for Diarrhoeal Disease Research, Bangladesh (icddr,b) for storage at ≤-70°C until analysis.

Venous blood samples were collected at the postnatal day 60 visit (or if necessary, rescheduled to a later date up to day 180 of age), and processed to serum at MCHTI or MFSTC[30], before transfer to icddr,b for storage at <-70C until analysis.

### Fecal iron quantification

The dry weight of each stool sample was recorded following sample processing as described for optimization and validation (Part A). Fecal iron concentration was measured using the AAS assay in the Immunobiology, Nutrition and Toxicology Laboratory (INTL) at icddr,b, as described above (Part A). For stool samples with absorbances below the lower limit of quantification (LLOQ) of 0.2mg per 100g dry stool, values were imputed at one-half the LLOQ (i.e., 0.1 mg per 100 g dry stool) (Section B Supplementary Material).

### Serum ferritin and C-reactive protein (CRP)

Serum ferritin was quantified using an automated chemiluminescence assay kits (007P65030, Abbott) on an Abbott Alinity analyzer. The between-and within-assay CVs were 5.82% and 2.78%, respectively, with a reportable range of 1.98 – 167.56ng/ml and an LLOQ of 1.98ng/ml. Serum CRP was measured using the high-sensitivity CRP assay on the Beckman Coulter autoanalyzer (OSR6199); the LLOQ was 0.08 mg/L and values below the LLOQ were imputed at 1/2 LLOQ (0.04 mg/L).

### Statistical analysis

The distributions of variables across intervention arms were visualized using histograms, k-density plots, and boxplots, as appropriate. Continuous normally and non-normally distributed variables were summarized using means (with standard deviations, SD) or medians (with 25^th^ and 75^th^ percentiles), respectively. Categorical variables were described as the frequencies and proportions in each category. Study outcomes (i.e., stool iron, serum ferritin, serum CRP) were natural log (ln)-transformed to approximate a normal distribution for regression analyses.

Average biomarker concentrations in each of the placebo and intervention groups were expressed as geometric means with 95% confidence intervals (CI).

Primary analyses were conducted using an intention-to-treat (ITT) approach, whereby participants who received the LP1 or LP1+FOS intervention were combined to create a single LP1 intervention group. Similarly, participants who received LP7 or LP7+FOS were combined to create a single LP7 intervention group. To examine the effect of probiotic administration (as 1-or 7-day regimens) on stool iron content and iron status, unadjusted linear regression models were fitted using intervention groups as the categorical exposure variable and the ln-transformed biomarker of interest as the continuous outcome measure. Effect estimates were back-transformed and reported as mean percent differences and 95%CIs for each intervention group relative to placebo. In further analyses, the combined intervention groups (1-or 7-day regimens) were disaggregated to examine the effect of each probiotic regimen (i.e., with or without FOS) relative to placebo, using the same modeling approach as in the primary analyses.

In supplementary analyses, we conducted subgroup analyses stratified by selected infant characteristics (sex, breastfeeding pattern up to 14 days of age, mode of delivery, and birthweight). We also examined these selected infant characteristics as effect-modifiers of the intervention effect on the primary outcome (stool iron concentration) by introducing interactions of the hypothesized modifiers with intervention group in the linear regression model, acknowledging these were exploratory analyses given the low sample sizes within each stratum. For this analysis, breastfeeding pattern up to 14 days postnatal age was dichotomized as exclusively breastfed (EBF) and not EBF, as most infants during the first 2 weeks after delivery received only human milk, resulting in a very small sample size in the other feeding-related sub-groups (e.g., partially, predominantly, or not breastfed). In a further supplementary analysis, separate unadjusted linear regression models were used to examine associations between the primary outcome (stool iron concentration) and each of the following covariates of interest: infant sex, breastfeeding pattern up to 14 days of age, mode of delivery and birthweight. The purpose of this additional exploratory analysis was to examine if the outcome measure captured meaningful between-infant variation in stool iron content and thereby corroborated the findings of the pilot testing of pooled samples (Part A). CRP was previously shown to be similar in the probiotic/synbiotic versus placebo groups in the primary report of the LP trial[30]; however, to address the potential contribution of systemic inflammation to increases in ferritin, we analyzed ln-transformed serum CRP overall and across intervention groups among infants included in this sub-study.

## Results

### Part A: Fecal iron assay optimization and validation

The optimized assay had a LLOQ of 0.2mg Fe/100g stool and LOD of 0.02 mg Fe/L (Section B in Supplementary Material). Accuracy using iron standards was 91 to 99% (Supplementary Table S2), and intra-run precision and inter-run coefficients of variation (CV%) were below 10% using reference materials and pooled stool samples (Supplementary Table S3). Recovery of iron from stool samples ranged from 93 to 112%, whereas recoveries for rice flour and turmeric powder were 98% and 99%, respectively (Supplementary Table S4).

Among 32 pools of infant stool samples, iron concentrations ranged from 0.1 to 12.35mg Fe/100g dry stool (Figure 1). Stool iron concentrations were generally higher at enrolment (0-4 days of age) than at later ages, but differences between samples collected at 10-18 days and 56-64 days of age were minimal (Supplementary Figure S2). Stool iron concentrations were lower in pools from infants who were exclusively or predominantly breastfed versus those partially breastfed (or who had not received breast milk) at postnatal age 0 to 4 days (Supplementary Figure S3a) and at 2 weeks and 2 months old (Supplementary Figure S3b). Stool iron concentration did not notably vary by sex (Supplementary Figure S4) nor neonatal underweight status (Supplementary Figure S5).

**Figure 1:**
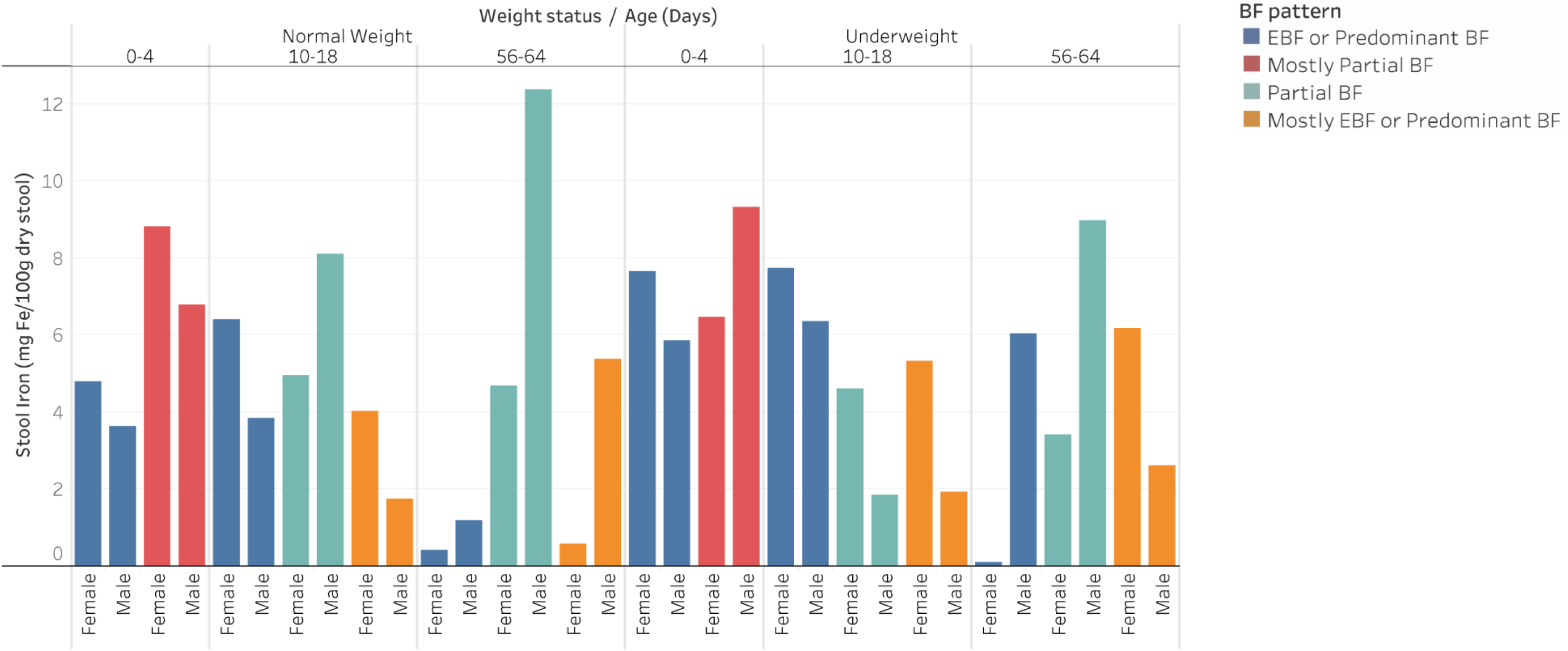
Stool iron concentrations in pooled samples categorized by sex, age, weight status, and breastfeeding pattern. N=32 pools. Each bar represents the measured stool iron concentration in one pooled sample. Pools are grouped by age and weight status, and breastfeeding patterns are distinguished by color. Pools originally designated as “Partial or No BF” are labelled as “mostly Partial BF” and “No BF” pools are labelled as “Mostly EBF + predominant BF” due to misclassification of the feeding status of some samples at the time the pools were generated (See Section C in Supplementary Material for further details).

Part B: Fecal iron quantification in a randomized controlled trial

Among 519 infants enrolled in the SEPSiS LP trial, 395 (76%) infants were included in this sub-study: 145 infants had only a day-14 stool sample, 88 infants had only a day-60 serum sample (actual sample collection ranged from day 60 to 194, whereby ∼91% of samples were collected by 3 months of age) and 162 infants had both a day-14 stool sample and a day-60 serum sample. In total, 307 participants contributed a stool iron measurement and 251 participants contributed a serum ferritin measurement (Figure 2). The actual timing of collection of stool samples ranged from 13 to 27 days postnatal age, with 93% collected from days 14 to 17 inclusive. Baseline characteristics were generally similar across the intervention groups (Table 1). No infants had a diarrheal event during the IP administration period and no infants were reported to have received any iron supplements during the first two weeks postnatal age. Nearly all infants (99%) received all 7 doses of their assigned intervention before the 14-day stool collection.

**Figure 2:**
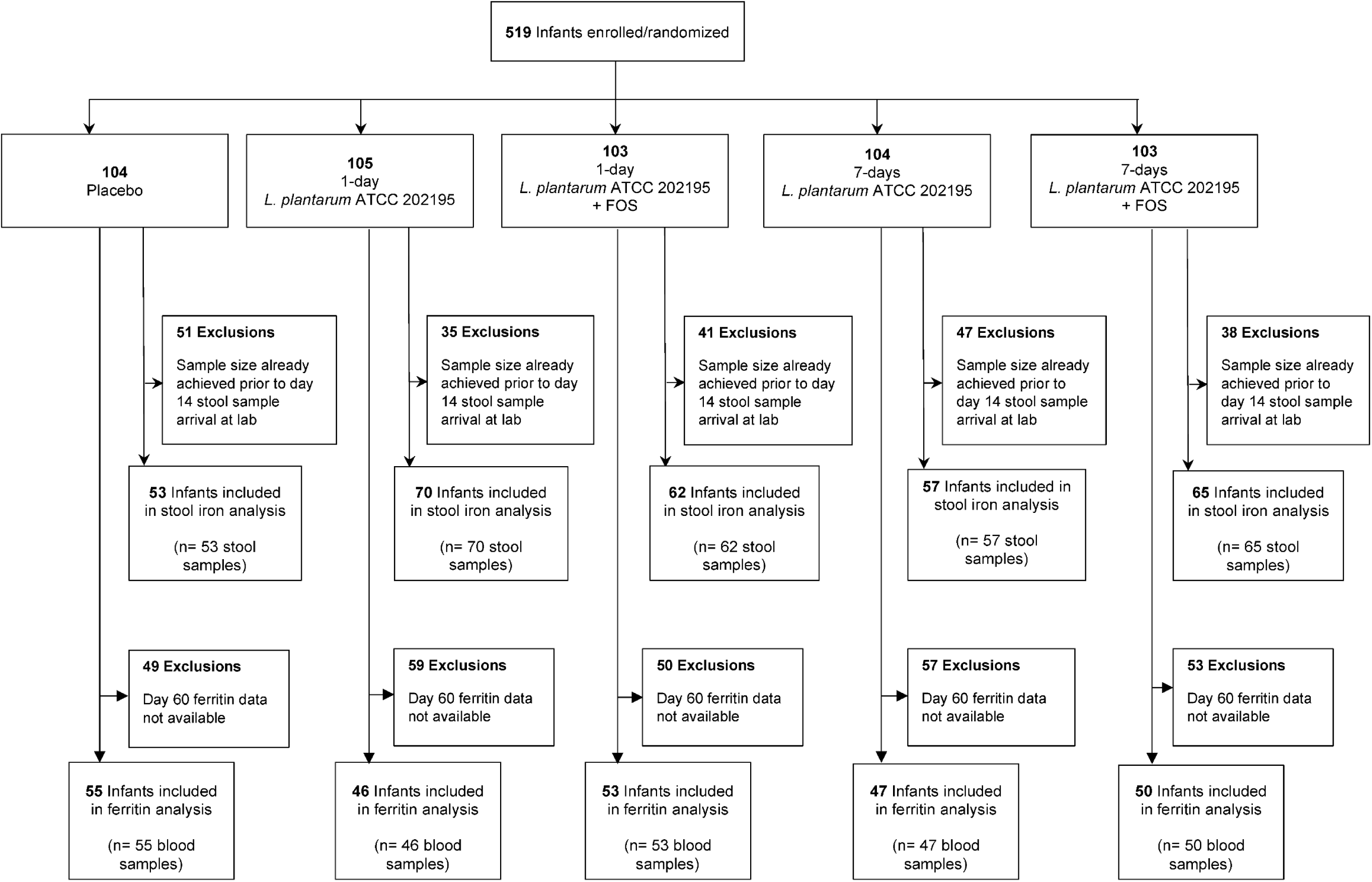
Study CONSORT Flow Diagram

**Table 1.**
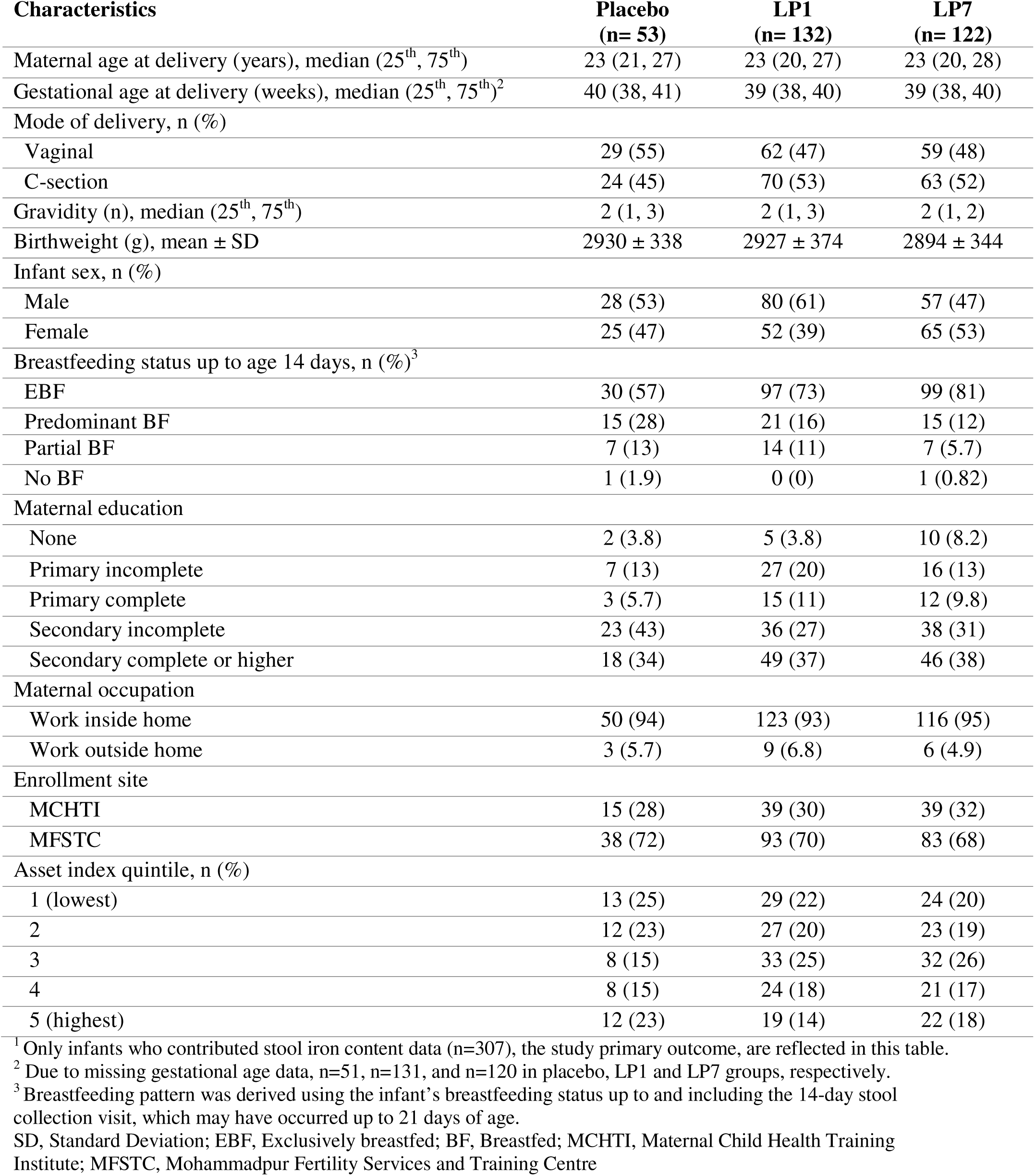
Participant characteristics, by intervention group (n=307)^1^.

| **Characteristics** | **Placebo**<br>**(n= 53)** | **LP1**<br>**(n= 132)** | **LP7**<br>**(n= 122)** |
| --- | --- | --- | --- |
| Maternal age at delivery (years), median (25<sup>th</sup>, 75<sup>th</sup>) | 23 (21, 27) | 23 (20, 27) | 23 (20, 28) |
| Gestational age at delivery (weeks), median (25<sup>th</sup>, 75<sup>th</sup>)<sup>2</sup> | 40 (38, 41) | 39 (38, 40) | 39 (38, 40) |
| Mode of delivery, n (%) |  |  |  |
| Vaginal | 29 (55) | 62 (47) | 59 (48) |
| C-section | 24 (45) | 70 (53) | 63 (52) |
| Gravidity (n), median (25<sup>th</sup>, 75<sup>th</sup>) | 2 (1, 3) | 2 (1, 3) | 2 (1, 2) |
| Birthweight (g), mean ± SD | 2930 ± 338 | 2927 ± 374 | 2894 ± 344 |
| Infant sex, n (%) |  |  |  |
| Male | 28 (53) | 80 (61) | 57 (47) |
| Female | 25 (47) | 52 (39) | 65 (53) |
| Breastfeeding status up to age 14 days, n (%)<sup>3</sup> |  |  |  |
| EBF | 30 (57) | 97 (73) | 99 (81) |
| Predominant BF | 15 (28) | 21 (16) | 15 (12) |
| Partial BF | 7 (13) | 14 (11) | 7 (5.7) |
| No BF | 1 (1.9) | 0 (0) | 1 (0.82) |
| Maternal education |  |  |  |
| None | 2 (3.8) | 5 (3.8) | 10 (8.2) |
| Primary incomplete | 7 (13) | 27 (20) | 16 (13) |
| Primary complete | 3 (5.7) | 15 (11) | 12 (9.8) |
| Secondary incomplete | 23 (43) | 36 (27) | 38 (31) |
| Secondary complete or higher | 18 (34) | 49 (37) | 46 (38) |
| Maternal occupation |  |  |  |
| Work inside home | 50 (94) | 123 (93) | 116 (95) |
| Work outside home | 3 (5.7) | 9 (6.8) | 6 (4.9) |
| Enrollment site |  |  |  |
| MCHTI | 15 (28) | 39 (30) | 39 (32) |
| MFSTC | 38 (72) | 93 (70) | 83 (68) |
| Asset index quintile, n (%) |  |  |  |
| 1 (lowest) | 13 (25) | 29 (22) | 24 (20) |
| 2 | 12 (23) | 27 (20) | 23 (19) |
| 3 | 8 (15) | 33 (25) | 32 (26) |
| 4 | 8 (15) | 24 (18) | 21 (17) |
| 5 (highest) | 12 (23) | 19 (14) | 22 (18) |

<sup>1</sup> Only infants who contributed stool iron content data (n=307), the study primary outcome, are reflected in this table.
<sup>2</sup> Due to missing gestational age data, n=51, n=131, and n=120 in placebo, LP1 and LP7 groups, respectively.
<sup>3</sup> Breastfeeding pattern was derived using the infant's breastfeeding status up to and including the 14-day stool collection visit, which may have occurred up to 21 days of age.
SD, Standard Deviation; EBF, Exclusively breastfed; BF, Breastfed; MCHTI, Maternal Child Health Training Institute; MFSTC, Mohammadpur Fertility Services and Training Centre

Among 307 participants with a fecal iron measurement in a day-14 stool sample (range: 0.1 to 95.6 mg Fe/100g dry stool), there was no effect of the 1-or 7-day regimens of LP202195 compared to the placebo group on fecal iron content (Table 2; Figure 3A). Among 251 participants with serum ferritin measured in a day-60 serum sample (range: 9.6 to 1219 ng/ml), there was no effect of 1-or 7-day regimens of LP202195 on serum ferritin concentrations at 2 months of age (Table 2; Figure 4). Inferences were unchanged in disaggregated analysis of groups based on FOS co-administration (i.e., with or without FOS), such that there were no effects of the intervention on either day-14 stool iron or day-60 serum ferritin concentrations in any intervention group compared to placebo (Table S6). Serum CRP was generally very low (median: 0.4 mg/L; 25^th^ and 75^th^ percentiles: 0.2 and 0.9 mg/L; n=251 infants). Only 10/251 infants (4%) had CRP values above 5 mg/L, and CRP did not differ across intervention groups (Supplemental Figure S6); therefore, no further analyses were conducted to consider effects of inflammation on between-group comparisons of serum ferritin.

**Figure 3.**
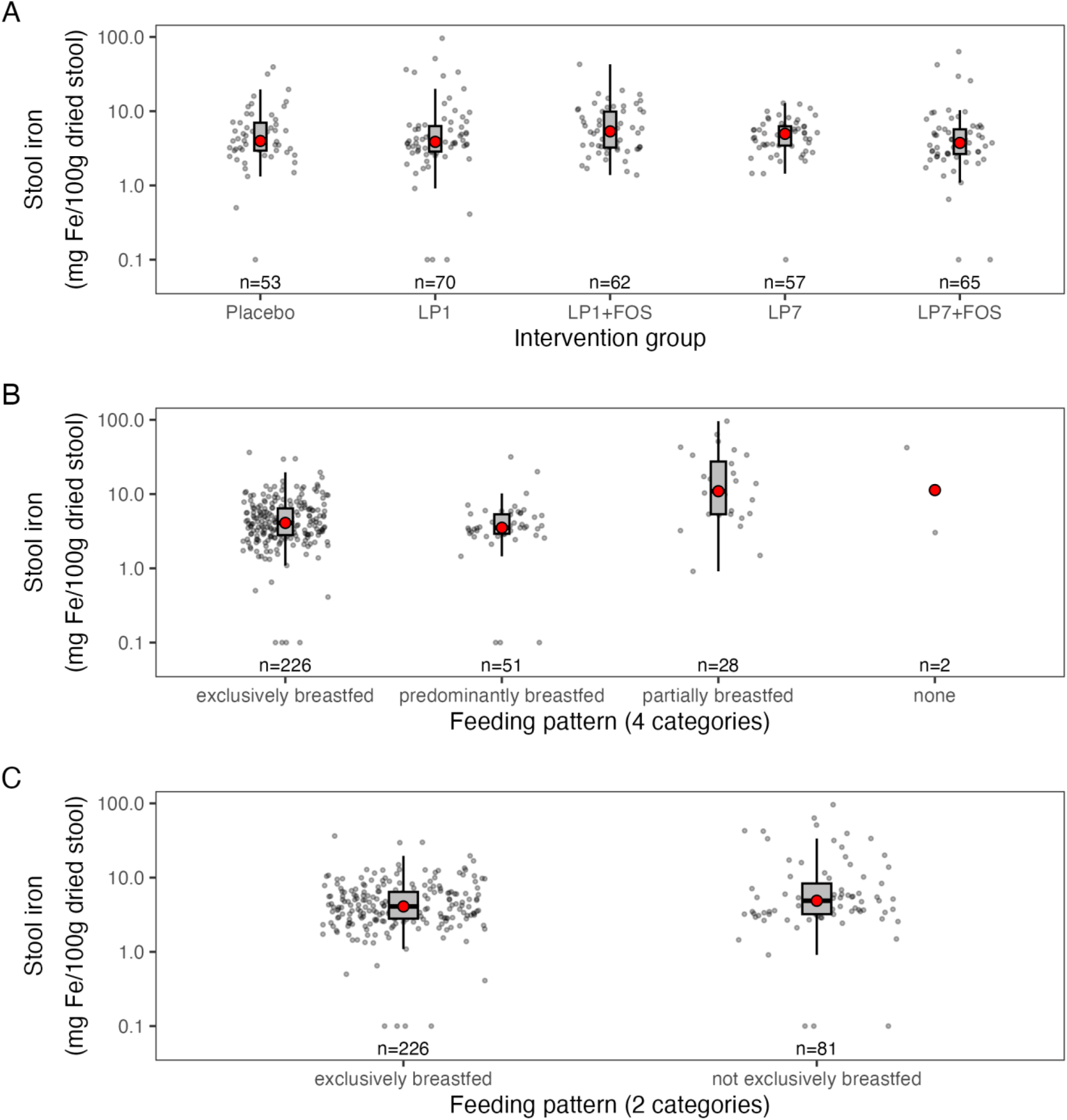
Stool iron concentration (mg Fe/100g dried stool), by intervention group (panel A) and feeding pattern as 4 categories (panel B) or 2 categories (panel. **C)**. (n=307 samples, one sample per infant). The grey boxplot represents the interquartile range, individual data points are shown as grey circles, and red circle denotes median. Y-axis shown as log scale. Differences in stool iron (natural log-transformed) across intervention groups (Panel A) or feeding patterns (Panel C) were estimated using linear regression model; a Wald test was conducted as a global test of any significant differences between any group and reference (Panel A: p-value = 0.2688 [intervention group, placebo as reference]; Panel C: p-value: 0.0286 [2 category feeding pattern, exclusively breastfed as reference]).

**Figure 4.**
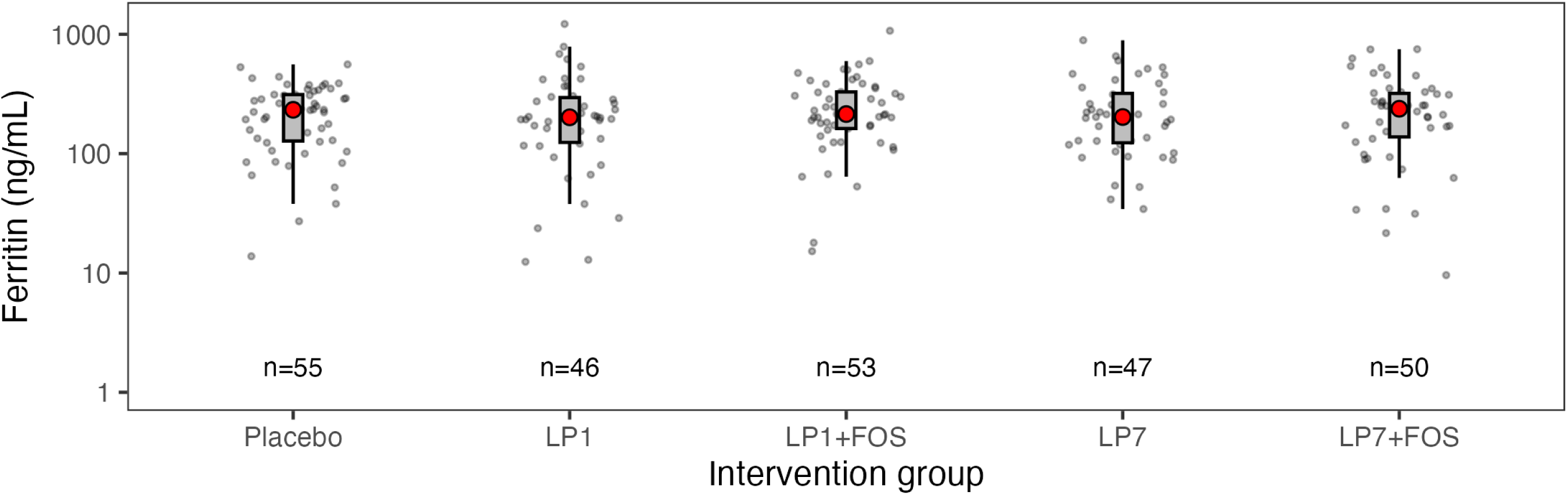
Serum ferritin concentrations (ng/mL), by intervention group. (n=251 samples, one sample per infant). The grey boxplot represents the interquartile range, individual data points are shown as grey circles, and red circle denotes median. Y-axis shown as log scale. Differences in ferritin (natural log-transformed) across intervention groups were estimated using linear regression model; a Wald test was conducted as a global test of any significant differences between any IP group and placebo (p-value = 0.9037).

**Table 2.** Effect of 1-and 7-day regimens of LP202195 (with and without FOS) on stool iron and serum ferritin concentrations^1^.

|  | Placebo |  | 1-Day Intervention |  |  | 7-Day Intervention |  |  |
| --- | --- | --- | --- | --- | --- | --- | --- | --- |
|  | n | Geometric Mean (95% CI) | n | % Difference (95% CI) | *p* | n | % Difference (95% CI) | *p* |
| **Stool Iron Concentration at 14 days of age (mg Fe/100g dry stool)** | 53 | 4.3 (3.3, 5.6) | 132 | 9.8 (-19, 49) | 0.54 | 122 | -6.1 (-31, 28) | 0.69 |
| **Serum Ferritin Concentration at 60 days of age (ng/mL)** | 55 | 189 (155, 232) | 99 | 2.4 (-22, 35) | 0.86 | 97 | 1 (-23, 33) | 0.94 |

<sup>1</sup> Effect estimates are reported as % mean differences (with 95% CI) relative to placebo, estimated using unadjusted linear regression models with intervention group as the categorical exposure variable.

There was no significant modification of the effects of the 1-or 7-day regimen of LP202195, with and without FOS, on infant stool iron concentration at 14 days postnatal age, in analyses in subgroups of participants stratified by sex, breastfeeding status, mode of delivery, and birthweight, and none of these factors significantly modified the intervention effect (Table 3).

**Table 3.**
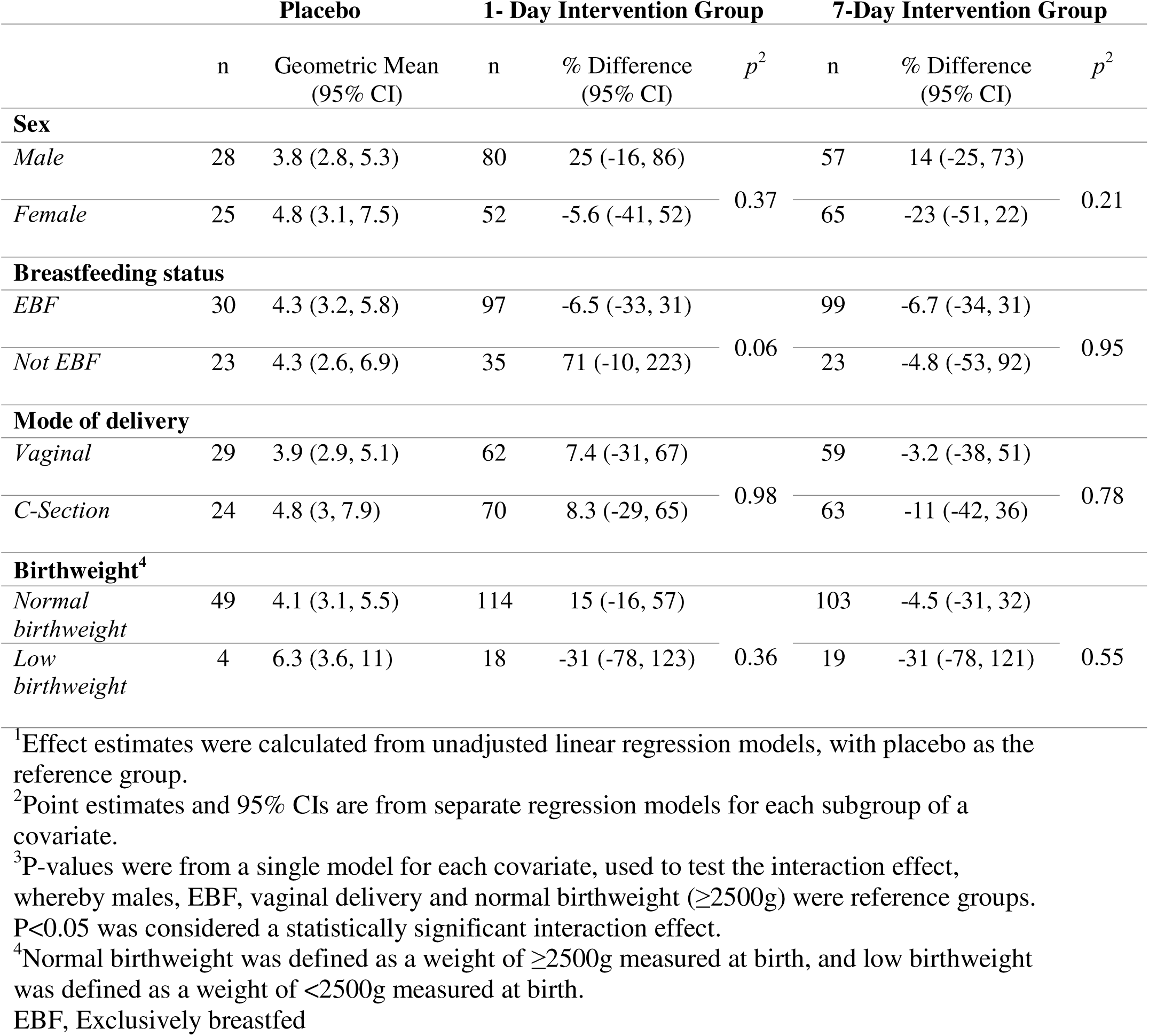
Effect of LP202195, with and without FOS, for 1-or 7-days, relative to placebo, on infant stool iron concentration at 14-days, by sex, breastfeeding status, mode of delivery or birthweight strata^1, 2, 3.^

In supplementary analyses, stool iron concentrations were significantly lower among infants who were exclusively breastfed compared to those who were not (Table 4; Figures 3B and 3C), but stool iron concentrations did not vary significantly by sex, mode of delivery, or birthweight in unadjusted analyses (Table 4).

**Table 4.** Associations of perinatal factors with stool iron concentration at 14 days of age.

| **Factor** | **N<sub>A</sub>;N<sub>B</sub><sup>1</sup>** | **% Difference**<br>**(95% CI)** | ***p*** |
| --- | --- | --- | --- |
| **Male (vs Female)** | 165;142 | 6.8 (-14, 32) | 0.55 |
| **EBF (vs not EBF)** | 226;81 | -23 (-40, -2.7) | 0.03 |
| **Vaginal delivery (vs C-section)** | 150;157 | -18 (-34, 1.4) | 0.066 |
| **Normal birthweight (vs low birthweight)** | 266;41 | -3.8 (-30, 32) | 0.81 |

<sup>1</sup>N<sub>A</sub>: sample size in primary sub-group; N<sub>B</sub>: sample size in the reference subgroup (identified in parentheses in column to the left)
EBF, Exclusively breastfed

Given the negative association between EBF and stool iron content, and acknowledging that EBF was not evenly distributed across the intervention groups, we conducted a covariate-adjusted model for the primary outcome with adjustment for EBF status at 14 days of age; this post-hoc analysis did not change the primary inferences such that there was no effect of the intervention on fecal iron following 1 day (% difference=15%, 95%CI:-15%, 56%; *P*=0.37) or 7 days (% difference=0.22%, 95%CI:-27%, 37%; *P*=0.99) of probiotic and/or synbiotic administration.

## Discussion

We optimized an optimized AAS assay for stool iron quantification, which had acceptable sensitivity, accuracy, precision, and recovery, and was feasibly applied to the determination of fecal iron concentration in samples from young infants in a cohort study and RCT in Bangladesh. We did not find evidence of an effect of LP202195, when administered with and/or without FOS for 1-or 7-days, on stool iron concentration at 14 days, and similarly there was no evidence of an effect on serum ferritin at 60 days postnatal age. Although the small sample size and early age timepoints at which samples were collected are important limitations of this trial sub-study, we demonstrated the practical considerations and challenges of developing and implementing a stool iron quantification assay for application in the context of a randomized trial of a probiotic/synbiotic in early infancy.

As an indirect biomarker of relative iron bioavailability, fecal iron has the advantage of being measurable in stool samples collected for other purposes in the context of microbiome studies and probiotic/synbiotic trials, in contrast to iron status biomarkers (which do not reflect intestinal iron dynamics, and require invasive blood sampling) and the relative complexity of stable-isotope studies[34]. However, analytical methods to measure stool iron are not widely used and there is no standard method in the literature, particularly for pediatric cohorts in which low stool sample volumes may fail to meet typical assay requirements[29]. Therefore, following development and validation of an AAS assay for infant stool aliquots by adapting previously published protocols, and achieving acceptable performance after assay optimization, the method was considered suitable for application in the context of a RCT.

The observed mean and range of stool iron concentration at 14 days postnatal age in the present study (4.3 mg Fe/100g dry stool, range of 0.1 to 95.6mg Fe/100g dry stool) were consistent with results previously reported for infants of similar ages in Chile and England[32, 33]. For example, Pizarro *et al*. reported that infants 3 to 15 months old in Chile had stool iron levels of 8.9mg Fe/100g stool when fed non-iron fortified formula and 27mg Fe/100g stool when iron-fortified formula was consumed[32]. Similarly, newborn infants aged 7 to 10 days in England who received iron-fortified formula had an average stool iron concentration of 9.2mg Fe/100g stool[33]. Numerous factors are known to affect stool iron concentration including diet, intestinal microbiota, inflammation, and intestinal sloughing and bleeding[29, 35], which may explain the wide range of stool iron concentrations among infants in the present study. Corroborating the preliminary findings using pooled samples in the pilot study, stool iron concentrations were significantly lower among infants in the RCT who were EBF within the first 14 days compared to those who were not EBF. This finding may be explained by the iron present in fortified infant formula, which is at higher concentrations but less bioavailable than the iron in human milk[4, 6]. When interaction effects were tested, none of the factors included in the analysis, including breastfeeding status, significantly modified the intervention effect, although we recognize the small sample size of the non-EBF group was a major limitation of the subgroup analyses. Nonetheless, the negative association between breastfeeding status and stool iron was in the direction expected, thereby supporting robustness of the outcome measure.

The LP202195 probiotic strain used in the present nested sub-study was selected based on the findings of Panigrahi *et al*., which demonstrated that this strain reduced the risk of neonatal sepsis in India[36]. However, effects of probiotics are species-and strain-specific[37]. To date, most evidence in support of the role of probiotics in improving iron absorption has stemmed from research using *L. plantarum* strain 299v, for which potential mechanisms of action include alteration of intestinal microbiota composition, production of metabolites that may reduce intestinal pH[21, 22] and increase iron solubility[23]. It is possible that the LP202195 strain does not substantially affect iron absorption, whereas findings may have differed with use of alternative strains (such as 299v). In the present trial, LP202195 stool abundance (cells per µg DNA or per g stool) was greater in the intervention arms (versus placebo) between ∼14 and 60 days of age, but progressively declined in the post-intervention period and did not persist beyond ∼2 months, indicating a lack of sustained colonization in this study population[30]. Therefore, a longer duration of probiotic administration to sustain high abundance may be necessary to produce measurable effects on iron status to 2 months of age and beyond. It is also possible that the iron absorption-promoting effects of LP202195 are only realized if it is co-administered with an exogenous iron source, as has been demonstrated in other studies[18, 20]. Hence, effects may be negligible during the early postnatal period when infants primarily consume human milk and their iron status is largely determined by prenatal maternal-fetal transfer, potentially explaining the null findings in the present study. However, two pediatric studies of lactobacilli intervention, conducted in comparatively older children, did not show improvements in iron status in the probiotic group, even when co-administered with extrinsic iron: among a population of pre-school children in Brazil with a high prevalence of anemia, serum ferritin decreased in response to a fermented milk beverage fortified with iron amino acid chelate and supplemented with *Lactobacillus acidophilus* but increased in children who received only the probiotic [27]; and, in predominantly iron sufficient 8-14 months old Thai children, there was no effect of synbiotic intervention with *Limosilactobacillus reuteri* and galactooligosaccharide on fractional iron absorption in response to single serving of a follow-up formula fortified with isotopically-labelled ferrous sulfate[28]. The present findings, alongside these two aforementioned studies, suggest a lack of benefit of lactobacilli-containing probiotics on iron uptake in young children. However, overall interpretation of any evidence of effect of probiotic/synbiotic administration on iron uptake and status in this age group is limited by heterogeneity in study designs, probiotic/synbiotic strains, outcome measurements and iron status prior to intervention.

We also measured serum ferritin at ∼2 months of age, whereby the mean serum ferritin concentration was 191ng/ml with a range of 9.6 to 1219ng/ml, consistent with the range of values observed among Swedish term-born infants in cord blood (8-1112ng/ml) and at 4 months of age (6-880ng/ml)[38]. Although fetal iron endowment primarily determines iron stores in early infancy, iron absorption is tightly regulated. Human milk has low concentrations yet highly bioavailable dietary iron, but when iron stores are sufficient (reflected by high ferritin), absorption is downregulated[6], limiting the potential for early probiotic interventions to meaningfully affect iron status.

An important limitation of quantifying fecal iron concentration is that it is not a direct measure of iron absorption; furthermore, sample processing (particularly lyophilization and ashing) was time-consuming and labor-intensive, and therefore the method may not be feasible for large cohorts or where resources are limited. In the present sub-study of a trial that was not primarily designed to examine stool iron and iron status, resources were available to analyze only one stool sample per infant, preventing an examination of stool iron trajectories with age. There were several other limitations of its application within the LP trial. First, the day-14 timepoint was selected for primary stool iron content analyses as most infants were expected to have completed the 7-day IP regimen by 14-days postnatal. However, the low but highly bioavailable iron content of human milk, as discussed above, likely contributed to the overall low stool iron levels observed, limiting any effect of the probiotic/synbiotic on either fecal iron or serum ferritin at such an early age. Data on maternal iron supplementation, maternal iron status, and cord blood ferritin concentration were not available but may have influenced infant iron stores; however, given the randomized design of the study, these factors are expected to have been evenly distributed across intervention groups and therefore we do not expect variation in baseline iron stores to have biased results of the primary analysis. In addition, the low sample size limited the inferences in sub-group analyses.

In conclusion, fecal iron quantification by AAS was valid and feasibly implemented in a trial of neonatal administration of *Lactiplantibacillus plantarum* ATCC 202195. A key advantage of the assay is that it only requires a relatively small aliquot of stored stool collected for other purposes, yet it is resource-intensive, does not quantify iron absorption, and may not be more informative than conventional measures of iron status (e.g., serum ferritin) when examining the effects of a probiotic/synbiotic intervention. In the present application of the assay in a RCT involving young infants, we did not find evidence that LP202195, when administered as either a probiotic or synbiotic regimen, influenced early measures of stool iron (2 weeks of age) or iron status (2 months of age). However, further studies are required to understand probiotic attributes in relation to host iron absorption and iron-gut microbiota interactions, whereby stable isotope techniques could potentially be considered for a direct reflection of iron absorption.

## Acknowledgements

This study was conducted as AZK’s doctoral research and therefore was reported in the PhD thesis. AZK and DER conceptualized the study; AZK, DER, RR, and AKR designed the study; RR, AKR and SA conducted laboratory analyses; AZK and HQ performed statistical analyses; AZK and LGP wrote the first draft of the paper; AZK, AKR, HQ, LGP, KMOC, SAS, AAM, RH, SA, SS, DER and RR edited and reviewed the paper. All authors have read and approved the final manuscript.

Investigational products were provided by International Flavors & Fragrances Inc. (IFF), but the company was not involved in the study design, implementation, data analysis, manuscript preparation, or publication decision. The authors are thankful to all SEPSiS project investigators and staff for their contributions to the design and implementation of the LP trial.

## Ethical Approval

This study was a sub-study of a randomized, placebo-controlled trial involving neonatal oral administration of LP202195, with or without FOS, for 1-or 7-days, which was approved by the ethical review committees at icddr,b (ERC protocol no. PR-20139), the Bangladesh Institute of Child Health, Dhaka Shishu Hospital, the ethical governing body for the Child Health Research Foundation (ERC protocol no. BICH-ERC-02-02-2019), the Directorate General of Drug Administration (DGDA), Bangladesh (DGDA/CTP-04/2016/672), and the Research Ethics Board at The Hospital for Sick Children, Toronto, Canada (REB no. 1000072200). Additionally, ethical approval was also obtained for this sub-study from the ethical review committees at icddr,b (ERC protocol no. PR-21099), and the Research Ethics Board at The Hospital for Sick Children, Toronto, Canada (REB no. 1000078635).

## Data Availability

De-identified data, code files, and all additional information required to reanalyze the data presented in this manuscript have been deposited at Borealis, The Canadian Dataverse Repository and will be made publicly available as of the date of publication. https://doi.org/10.5683/SP3/RJUT53

## List of Abbreviations

AAS: Atomic absorption spectrometry
ATCC: American type culture collection
CBL: Clinical Biochemistry Lab
CI: Confidence interval
EBF: Exclusively breastfed/Exclusive breastfeeding
FOS: Fructooligosaccharide
icddr,b: International Centre for Diarrhoeal Disease Research, Bangladesh
ID: Iron deficiency
IDA: Iron deficiency anemia
IP: Investigational product
INTL: Immunobiology, Nutrition and Toxicology Laboratory (INTL)
LMIC: Low-or middle-income country
LP1: *Lactiplantibacillus plantarum* ATCC 202195 without FOS for 1 day
LP1+FOS: *Lactiplantibacillus plantarum* ATCC 202195 with FOS for 1 day
LP202195: *Lactiplantibacillus plantarum* ATCC 202195
LP7: *Lactiplantibacillus plantarum* ATCC 202195 without FOS for 7 days
LP7+FOS: *Lactiplantibacillus plantarum* ATCC 202195 with FOS for 7 days
MCHTI: Maternal and Child Health Training Institute
MFSTC: Mohammadpur Fertility Services and Training Centre
SD: Standard deviation
SGA: Small for gestational age
WHO: World Health Organization

