## Supplementary Material for "Fecal iron quantification in a randomized controlled trial of *Lactiplantibacillus plantarum* ATCC 202195 in newborns in Dhaka, Bangladesh"

### A. SEPSiS study eligibility criteria

Inclusion and exclusion criteria for the SEPSiS observational study and SEPSiS LP trial were:

*Inclusion Criteria*

i. Infants up to and including four days of age
ii. Infant delivered at a study hospital
iii. Orally feeding currently
iv. Informed consent by parent or guardian
    a. Intends to maintain residence within the defined catchment areas (upon discharge from hospital) until 60 days of age

*Exclusion Criteria*

i. Birthweight < 1500 grams
ii. Death or major surgery considered to be highly probably within first week of life
iii. Major congenital anomaly of the gastrointestinal tract
iv. Maternal HIV infection and/or history of mother ever receiving anti-retroviral drug(s) for presumed HIV infection
v. Current mechanical ventilation and/or cardiac support (e.g., inotropes) and/or administration/prescription of parenteral antibiotics
vi. Any prenatal or postpartum use of non-dietary probiotic supplement by mother (during current pregnancy)
vii. Any postpartum use of non-dietary probiotic or prebiotic supplements to infant
viii. Current participation of the infant in another clinical trial
ix. Resides in the same household as another infant previously enrolled in the study, or any study within the research platform, who is currently <60 days of age; however, twins/multiples may all be enrolled simultaneously in this trial
x. Multiple gestation for which the number of liveborn infants from the same pregnancy exceeds two (i.e., triplets or higher order multiples)

### B. Optimization and validation of fecal iron quantification assay

**Table S1. Reference materials used for optimization of stool iron assay**

| **Sample** | **Composition** | **Source** | **Reconstitution Volume/Amount[1]** |
|---|---|---|---|
| Iron standard (CRM) | TraceCERT®, 1000 mg/L Fe in nitric acid | Sigma-Aldrich | 5 dilutions (~2mL) |
| Iron standard (CRM) | 5ppm Fe in nitric acid | Roche Diagnostics | ~1mL |
| NIST (SRM)[2] | Rice flour | NIST | ~0.2g |
| Rice flour[2] | Rice flour | Commercially procured | ~1g |
| Turmeric powder[2] | Turmeric powder | Commercially procured | 0.5g |
| Spiked sample | Spiked rice flour with known standard (CRM) | - | ~1g |
| Spiked sample | Spiked turmeric powder with known standard (CRM) | - | ~0.5g |
| Spiked sample | Spiked stool sample with known standard (CRM) | - | ~0.3g |

[1]Amounts shown in g are lyophilized weights, which varied by 0.05g. Final concentrations were calculated using weights reported to 2 decimal places.
[2] NIST, rice flour and turmeric powder were lyophilized in the same manner as the stool samples.

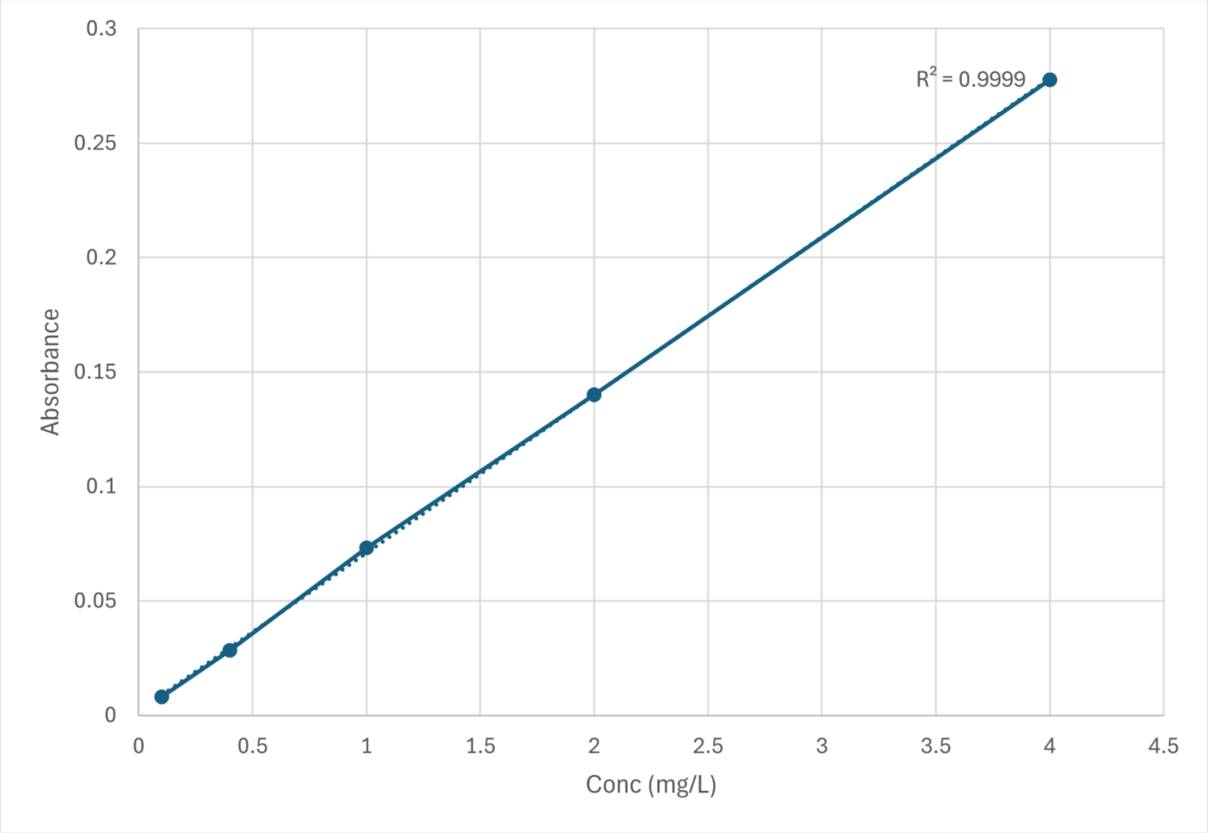

**Figure S1**. **Example of a standard curve for stool iron quantification using atomic absorption spectrometry (AAS).** Solid lines connect each plotted value. The dotted linear fit line almost entirely overlaps with the solid connecting lines. Absorbances were converted to iron concentrations (mg/L) using the standard curve, and stool iron concentration expressed in mg per kg of the lyophilized source material was derived using the following equation:

$$Stool\ iron\ (mg\ per\ kg\ dry\ weight) = Fe\ concentration\ \left(\frac{mg}{L}\right) \times \frac{Volume\ of\ ash\ solution\ (L)}{Dry\ sample\ weight\ (kg)} \times Dilution\ factor$$

Ash solution volume was usually 2mL of 0.2N HCl, and dilutions were usually 5-, 10-, 20-, or 40-fold (i.e., increased until an absorbance value in the reportable range was obtained).

For lyophilized stool samples, values from the equation above were multiplied by 0.1 to express stool iron concentrations in mg per 100 g dry stool.

**Lower limit of quantification (LLOQ):**

The LLOQ, also known as functional sensitivity[1], is the smallest concentration of an analyte that can be reliably quantified, which corresponded to the lowest concentration of the standard used in the calibration curve (0.4 mg Fe/L). To establish a fixed LLOQ expressed in the reporting units used for stool samples (mg Fe per 100 g dry stool), we applied the LLOQ to a nominal sample of 0.5 g lyophilized stool dissolved in 2 mL ash solution and diluted using the lowest dilution factor:

$$LOQ = 0.1\, mg\ Fe/L\ \times \frac{0.002\ L}{0.5\ g\ stool} \times\ 5\ \times\ 100 = 0.2\, mg\ Fe\ per\ 100\ g\ stool$$

**Lower limit of detection (LOD)**:

The LOD, also known as analytical sensitivity, is the smallest concentration at which the instrument can differentiate the analyte from the blank, using the following formula [2, 3]:

$$LOD = \frac{3.3\ \times SD\ of\ blank\ measurements}{Slope\ of\ standard\ curve}$$

The LOD was calculated as 0.02 mg Fe/L, based on a slope of the standard curve of 0.089 OD units/(mg/L) and the standard deviation (SD) of ten blanks of 0.0006 OD units.

**Accuracy**:

Accuracy of the method was verified by comparing results obtained from measurements of standards and reference materials from NIST (National Institute of Standards and Technology), with their pre-established values, to understand how closely the observed value approximated the true value. Accuracy was derived using the error rate equation shown below:

$$Accuracy = \left[1 - \left(\frac{Observed\ value - True\ Value}{True\ Value}\right)\right] \times 100$$

From repeated analysis of iron standards from Sigma Aldrich and Roche Diagnostics, and standard reference material from NIST (1894a and 1869a), accuracy was determined to be >90% (Table S2).

**Table S2. Accuracy of stool iron quantification assay using several reference materials**

| Run | Sample Name | N replicates | Min, max (mg/kg) | Mean ± SD iron concentration (mg/kg) | Accuracy |
|---|---|---|---|---|---|
| 1 | Standard (2ppm) Fe Standard for AAS, TraceCERT (CRM), Sigma-Aldrich | 1 | 1.93 | - | 97% |
| 2 | Standard (10ppm) Fe Standard for AAS, TraceCERT (CRM), Sigma-Aldrich | 4 | 10.38, 11.83 | 10.91 ± 0.697 | 91% |
| | Standard (5ppm) Fe Standard, Roche Diagnostics | 3 | 4.73, 5.05 | 4.93 ± 0.173 | 99% |
| 3 | Standard (10ppm) Fe Standard for AAS, TraceCERT (CRM), Sigma-Aldrich | 2 | 10.72, 11.09 | 10.91 ± 0.262 | 91% |
| | Standard (5ppm) Fe Standard, Roche Diagnostics | 1 | 4.93 | - | 99% |
| | NIST Standard Reference Material 1849a (175.6±29) | 4 | 175.17, 203.41 | 188.53 ± 14.4 | 93% |
| | NIST Standard Reference Material 1869a (164.7±3.7) | 4 | 165.86, 169.56 | 167.80 ± 1.52 | 98% |

**Precision**:

To calculate intra-run precision, replicates of standards, reference materials from NIST, pooled stool samples and internal QC samples (rice flour and turmeric powder) were measured, and the corresponding CV was calculated. Turmeric powder and rice flour were processed in the same manner as stool, whereby the powders were first made into a paste, and then lyophilized and ashed. Inter-run precision was also calculated using results from pooled samples and rice flour and corresponding CVs were calculated (Table S3).

**Table S3. Precision of iron content measurements in samples and reference materials**

| Sample Name | N replicates | Min, max (mg/kg) | Mean ±SD iron concentration (mg/kg) | Intra-run CV | Inter-run CV |
|---|---|---|---|---|---|
| Fe Standard (10ppm), TraceCERT (CRM), Sigma-Adrich | 4 | 10.35, 11.83 | 10.91 ± 0.697 | 6.4% | - |
| Fe Standard (5ppm), Roche Diagnostics | 3 | 4.73, 5.05 | 4.93 ± 0.173 | 3.5% | - |
| NIST Standard Reference Material 1849a (175.6±29) | 4 | 175.17, 203.41 | 188.53 ± 14.4 | 7.6% | - |
| NIST Standard Reference Material 1869a (164.7±3.7) | 4 | 165.86, 169.56 | 167.80 ± 1.52 | 0.9% | - |
| Pooled stool (Run 1) | 10 | 82.18, 100.26 | 89.41 ± 6.38 | 7.1% | 4.1% |
| Pooled stool (Run 2) | 2 | 87.19, 88.40 | 87.80 ± 0.86 | 1.0% | |
| Pooled stool (Run 3) | 2 | 89.65, 90.68 | 90.16 ± 0.72 | 0.8% | |
| Pooled stool (Run 4) | 2 | 90.88, 91.35 | 91.12 ± 0.33 | 0.4% | |
| Pooled stool (Run 5) | 2 | 97.18, 98.00 | 97.59 ± 0.58 | 0.60% | |
| Rice flour (Run 1) | 10 | 17.35, 21.11 | 19.75 ± 1.35 | 6.9% | 9.9% |
| Rice flour (Run 2) | 2 | 21.09, 21.90 | 21.49 ± 0.57 | 2.7% | |
| Rice flour (Run 3) | 2 | 17.59, 17.73 | 17.66 ± 0.10 | 0.6% | |

| | | | | | |
|---|---|---|---|---|---|
| Rice flour (Run 4) | 2 | 22.03, 22.19 | 22.11 ± 0.11 | 0.5% | |
| Turmeric powder | 6 | 114.81, 124.39 | 117.6 ± 3.70 | 3.1% | - |

**Recovery**:

To measure recovery, stool samples, rice flour and turmeric powder were spiked with a known concentration of iron standard (TraceCERT®, Sigma-Aldrich). Recovery of iron from stool samples was calculated as the percentage of the measured spike of the sample relative to the measured spike of the standard, which was added to the sample (Table S4).

**Table S4. Recovery measurements for iron concentrations**

| Sample Name | Expected Iron (mg/kg) | Measured iron (mg/kg) | Recovery |
|---|---|---|---|
| Rice flour spiked with 10ppm standard | 20.85 | 20.50 | 98% |
| Turmeric spiked with 10ppm standard | 131.69 | 129.85 | 99% |
| Stool sample spiked with 2ml 10ppm standard | 166.11 | 154.70 | 93% |
| Stool sample spiked with 2ml 10ppm standard | 166.11 | 159.10 | 96% |
| Stool sample spiked with 2ml 10ppm standard | 91.56 | 102.54 | 112% |
| Stool sample spiked with 2ml 10ppm standard | 91.56 | 102.32 | 112% |
| Stool sample spiked with 2ml 10ppm standard | 127.17 | 124.19 | 98% |
| Stool sample spiked with 2ml 10ppm standard | 127.17 | 122.60 | 96% |

#### C. Sample classification and pooling in the pilot demonstration of the fecal iron quantification assay

The stool iron quantification assay was applied to the analysis of pooled stool samples classified according to four infant characteristics: age, sex, infant feeding pattern, and neonatal underweight status.

Infant feeding pattern was based on feeding status up to the visit at which the stool sample was collected. The categories of feeding pattern defined as exclusively breastfed (EBF) or predominantly breastfed, partially breastfed and not breastfed, were based on definitions set by WHO[4] (outlined below).

Breastfeeding pattern definitions according to the WHO:

i. Exclusively breastfed (EBF): Infant consumed only human milk
ii. Predominantly breastfed: Infant consumed human milk with water, sugar water, honey, or other non-milk or non-formula liquid
iii. Partially breastfed: Sub-categorized based on the type of human milk substitute used (e.g., formula or other complementary food)
iv. None: Infant did not consume human milk

Categories i and ii were collapsed into a single category. Categories iii and iv were considered separately except for the birth time point (days 0-4), at which they were collapsed into one category because consumption of anything other than human milk was rare at such an early age. The categorization described here separated human milk from any formula or animal milk sources, due to their potential effect on stool iron.

Neonatal nutritional status, which was defined by whether infants were underweight, was based on the first available weight measurement by study personnel at enrolment (0-4 days of age).

Underweight was defined as <10$^{th}$ percentile of the healthy reference population (ie., weight-for-age z-scores <1.2), using WHO standards.  Since reported birthweight was not used, this cut-off was used as a proxy for small for gestational age (SGA), as WHO standards do not account for gestational age.

A total of 158 samples were grouped into 8 pools for the early age group and 12 for each of the later age groups, leading to a total of 32 pooled samples (Table S5).

Although the original intention was to include a Not breastfed (No BF) category in this study, and that is how samples were selected for pooling as shown in Table S5, the infant feeding status of several samples included in the No BF and partial/no BF pools was found to have been misclassified at the time of sample selection. Upon reclassification of infant feeding status, 80% of the samples at day 14 and day 60 that were originally classified as no BF were classified as EBF or predominantly breastfed ('EBF + predom BF') and a further 10% were classified as partial BF (i.e., overall 90% of samples in the no BF pools were re-classified), and all of the no BF pools included at least one sample for which feeding status was re-classified. Among day 0-4 samples, 6/18 samples (33%) originally classified as partial/no BF were re-classified as EBF + predom BF, and 3 of 4 pools labeled as partial/no BF included these re-classified samples. For transparency in reporting the findings related to feeding status, the strata are considered separately as originally selected, but the no BF category (ages 14 days and 60 days) has been re-labelled as "mostly EBF + predominant BF", and the partial + No BF category (day 0 samples) has been re-labelled as "mostly partial BF", given that most of the samples in these pools (12/18) were from infants classified as partially breastfed and none of the 18 samples were from non-breastfed infants following reclassification.

### Table S5. Pooling strategy for stool samples from the SEPSiS observational cohort

| Age | Feeding pattern | Sex | Neonatal underweight | Pool |
|---|---|---|---|---|
| Group A<br>Day 0-Day 4<br>(8 pools of 5 samples each= 40 samples) | EBF + predom BF | Male | Yes | A1 |
| | EBF + predom BF | Male | No | A2 |
| | EBF + predom BF | Female | Yes | A3 |
| | EBF + predom BF | Female | No | A4 |
| | Partial BF + No BF[1] | Male | Yes | A5 |
| | Partial BF + No BF[1] | Male | No | A6 |
| | Partial BF + No BF[1] | Female | Yes | A7 |
| | Partial BF + No BF[1] | Female | No | A8 |
| Group B<br>Day 10-Day 18<br>(12 pools of 5 samples each= 60 samples) | EBF + predom BF | Male | Yes | B1 |
| | EBF + predom BF | Male | No | B2 |
| | EBF + predom BF | Female | Yes | B3 |
| | EBF + predom BF | Female | No | B4 |
| | Partial BF | Male | Yes | B5 |
| | Partial BF | Male | No | B6 |
| | Partial BF | Female | Yes | B7 |
| | Partial BF | Female | No | B8 |
| | No BF[1] | Male | Yes | B9 |
| | No BF[1] | Male | No | B10 |
| | No BF[1] | Female | Yes | B11 |
| | No BF[1] | Female | No | B12 |
| Group C<br>Day 56-Day 64<br>(12 pools of 5 samples each= 60 samples) | EBF + predom BF | Male | Yes | C1 |
| | EBF + predom BF | Male | No | C2 |
| | EBF + predom BF | Female | Yes | C3 |
| | EBF + predom BF | Female | No | C4 |
| | Partial BF | Male | Yes | C5 |
| | Partial BF | Male | No | C6 |
| | Partial BF | Female | Yes | C7 |
| | Partial BF | Female | No | C8 |
| | No BF[1] | Male | Yes | C9 |
| | No BF[1] | Male | No | C10 |
| | No BF[1] | Female | Yes | C11 |
| | No BF[1] | Female | No | C12 |

[1]At the time of sample selection, samples were classified as "No BF"; however, these pooled samples were later recognized to include samples primarily collected from infants classified as EBF + predominant BF.

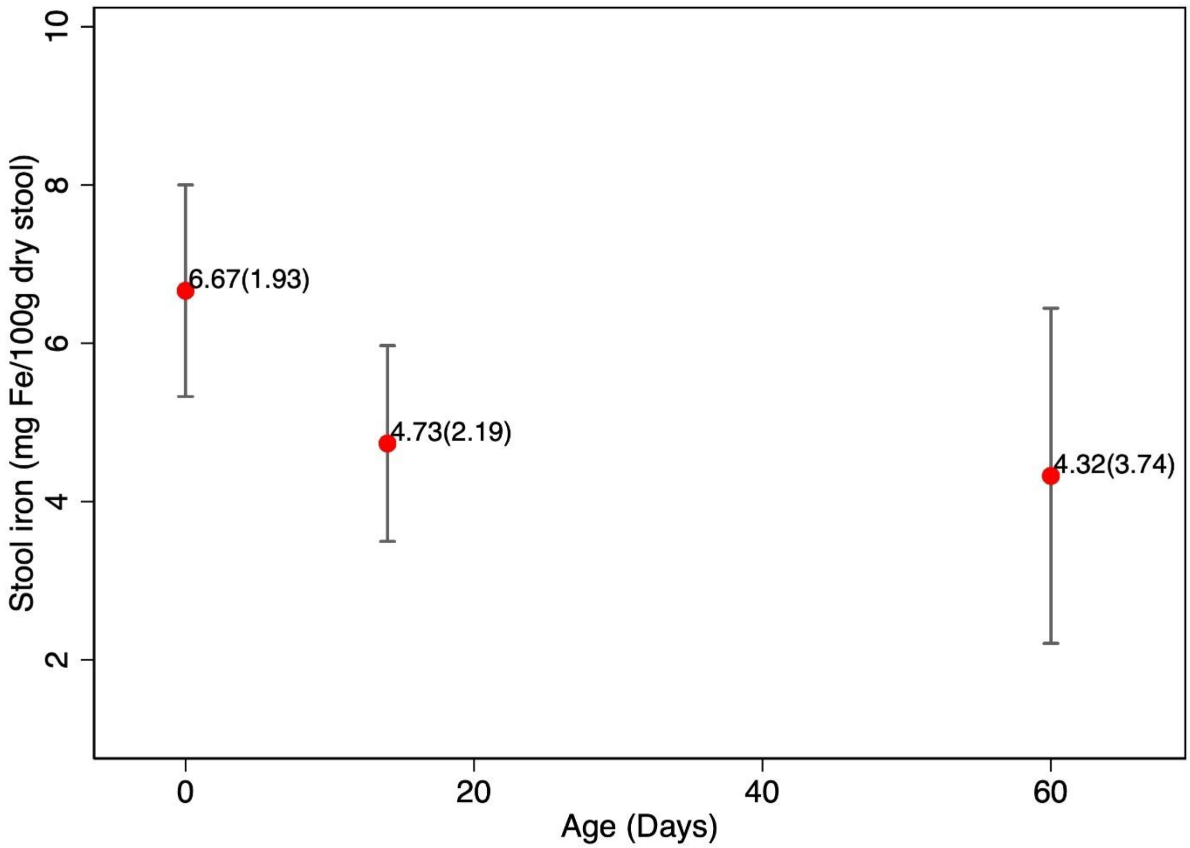

**Figure S2: Stool iron concentration at 0-4 days (8 pools), ~14 days (12 pools) and ~60 days of age (12 pools).** Data points indicate mean (SD) and the bar represents ±95% CI.

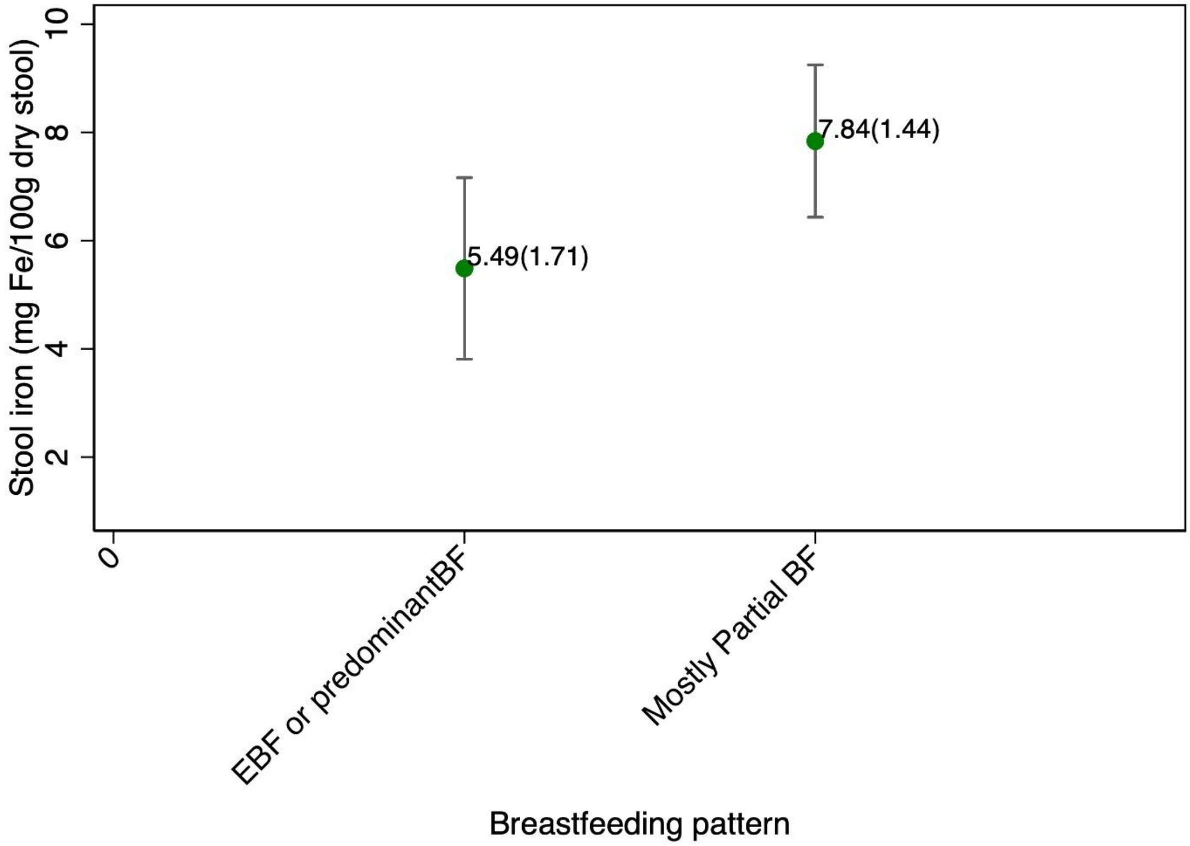

**Figure S3a: Stool iron concentration at 0-4 days of age, by infant breastfeeding pattern.** N=4 pools for each category. Data points indicate mean (SD) and the bar represents the 95% CI. Pools originally designated as "Partial or No BF" pools are labelled as "mostly Partial BF" due to feeding status misclassification

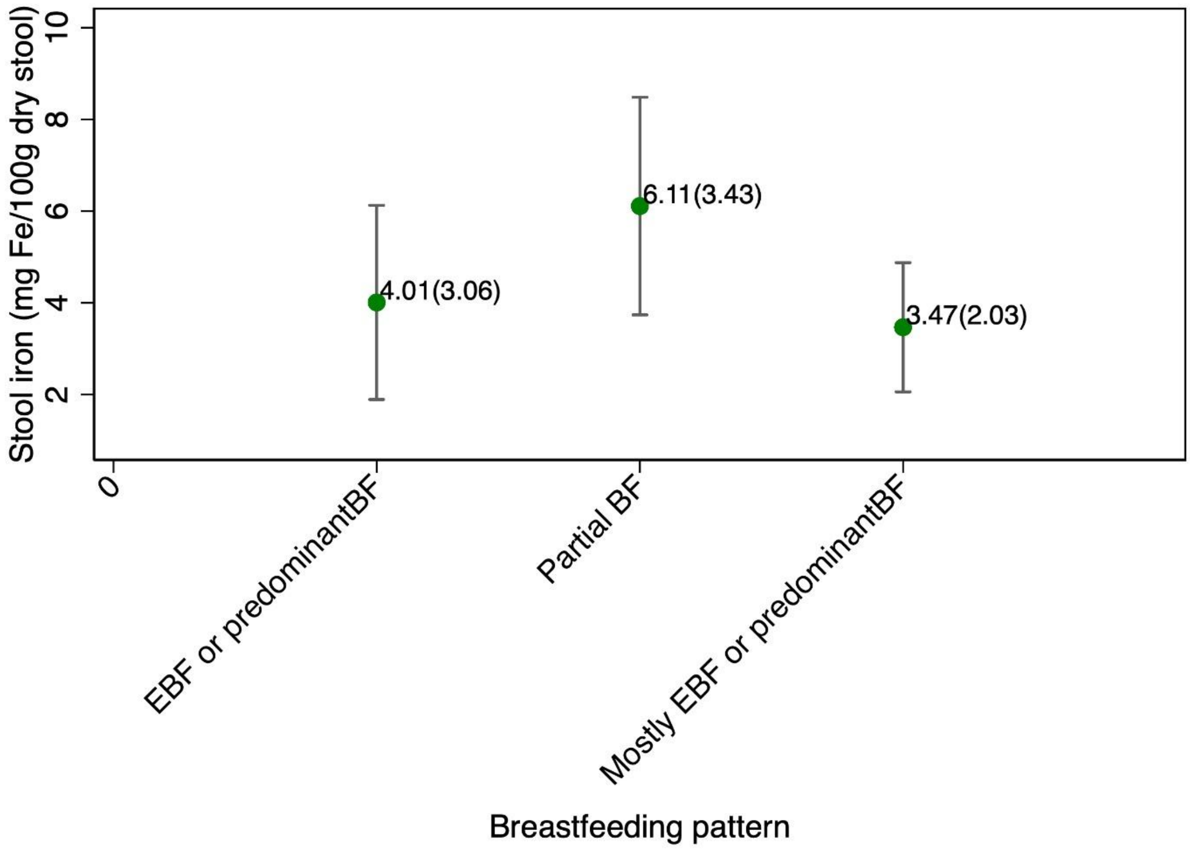

**Figure S3b: Stool iron concentration at 14 and 60 days of age, by infant breastfeeding pattern**. N=8 pools for each category. Data points indicate mean (SD) and the bar represents the 95% CI. Pools originally designated as “No BF” pools are labelled as “Mostly EBF + predominant BF” due to sample misclassification

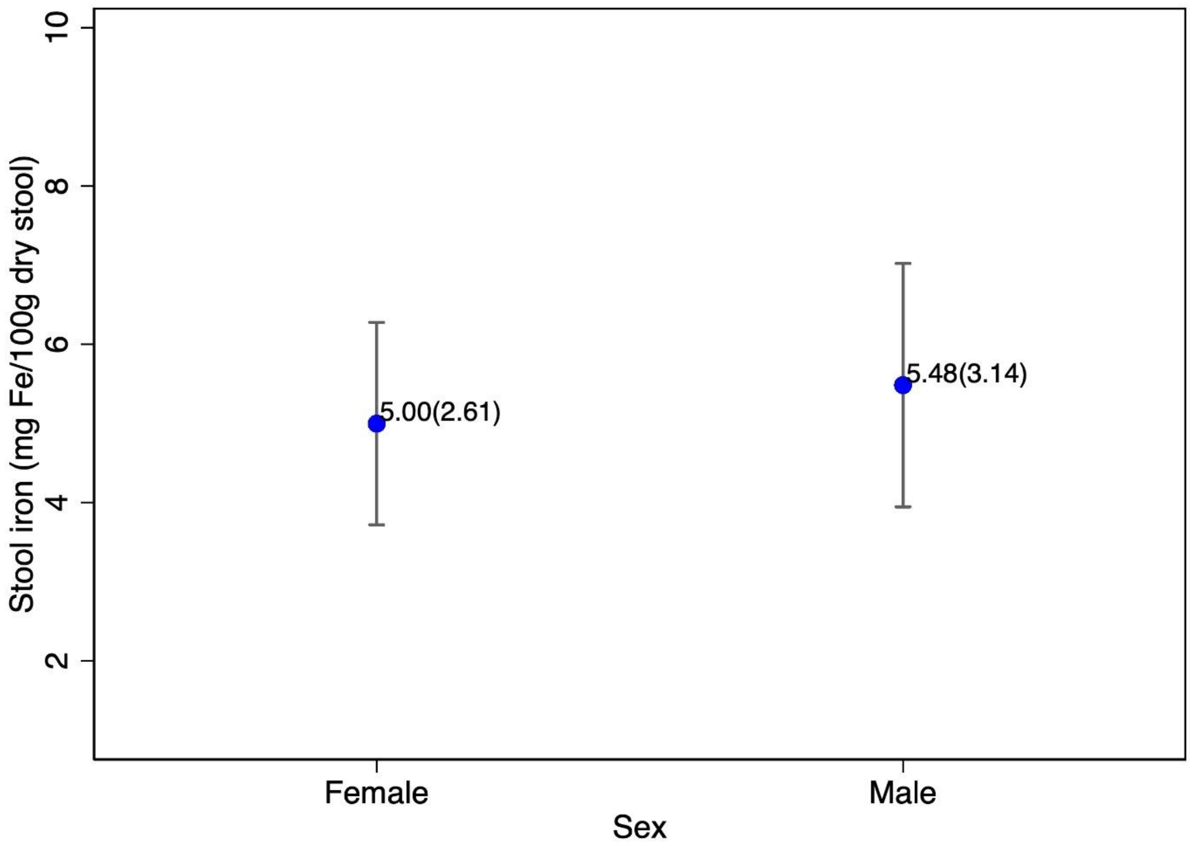

**Figure S4: Stool iron concentration, by infant sex.** N=16 pools in each category. Data points indicate mean (SD) and the bar represents the 95% CI.

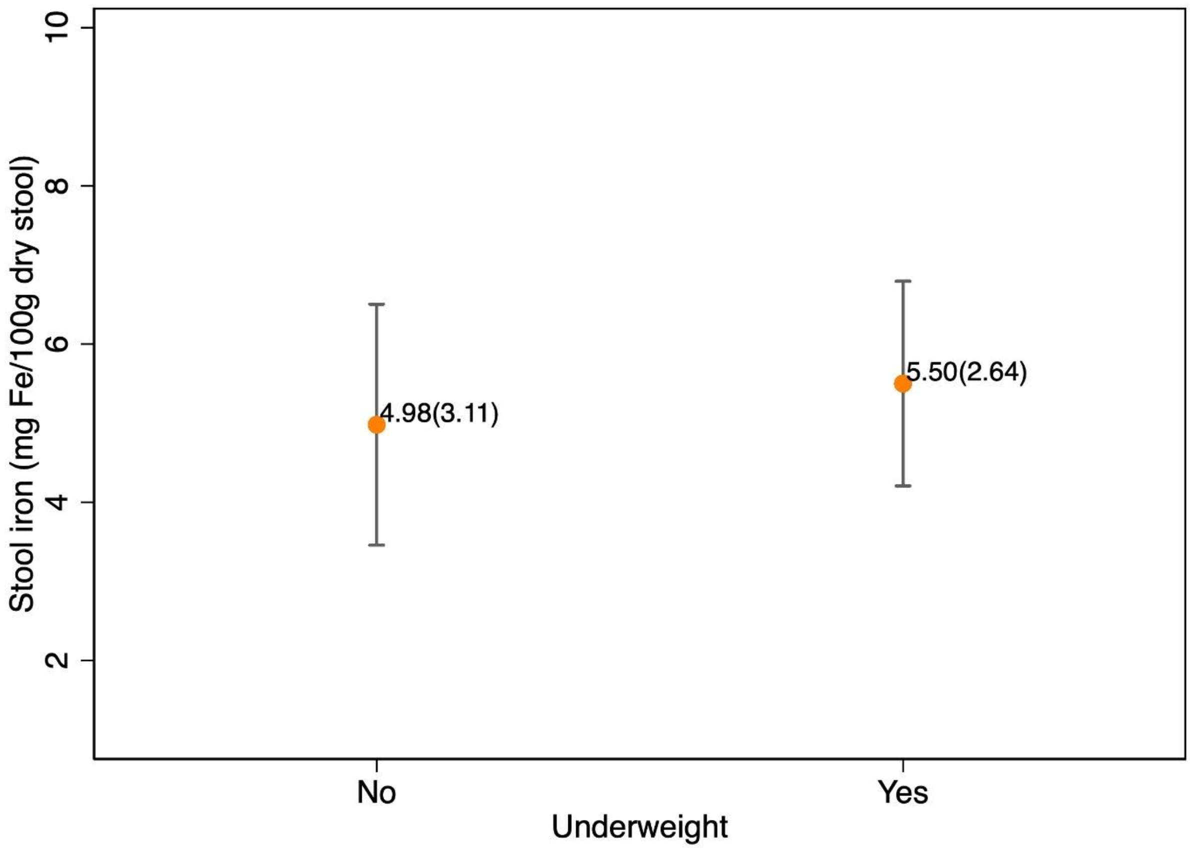

**Figure S5: Stool iron concentration, by infant nutritional (underweight) status.** N=16 pools in each category. Data points indicate mean (SD) and the bar represents the 95% CI.

#### D. Serum C-reactive protein (CRP)

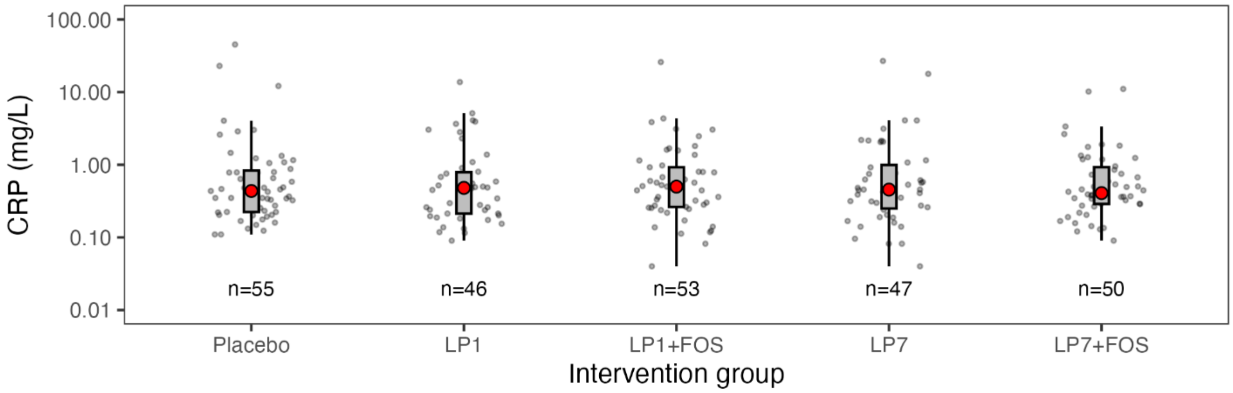

**Figure S6: High-sensitivity C-reactive protein (CRP) concentration (mg/L) in serum, by intervention group.** (n=251 samples, one sample per infant). Grey boxplot represents the interquartile range, individual data points are shown as grey circles, and the red circle denotes median. Y-axis shown as log-scale. For two samples with CRP values below the assay limit of detection (0.08 mg/L), values were imputed as 0.04. Differences in ln-transformed CRP across intervention groups were estimated using linear regression model; a Wald test was conducted as a global test of any significant differences between any IP group and placebo (p-value = 0.9974).

### E. Effect of L. plantarum 202195 on iron indicators, with or without FOS co-administration

**Table S6. Effect of *L. plantarum* 202195 and FOS on fecal iron and serum ferritin concentrations**

| | **1-Day LP** | | | **1-Day LP + FOS** | | | **7-Day LP** | | | **7-Day LP + FOS** | | |
|---|---|---|---|---|---|---|---|---|---|---|---|---|
| | ***n*** | ***% Difference (95% CI)*** | ***P*** | ***n*** | ***% Difference (95% CI)*** | ***P*** | ***n*** | ***% Difference (95% CI)*** | ***P*** | ***n*** | ***% Difference (95% CI)*** | ***P*** |
| **Stool Iron** | 70 | -3.4 (-31, 36) | 0.842 | 62 | 27 (-10, 80) | 0.18 | 57 | 1.3 (-29, 45) | 0.941 | 65 | -12 (-38, 24) | 0.456 |
| **Serum Ferritin** | 46 | -6.1 (-32, 30) | 0.704 | 53 | 10 (-19, 51) | 0.534 | 47 | 3.3 (-25, 43) | 0.845 | 50 | -1 (-28, 36) | 0.95 |

Results are reported as mean % difference (with 95% CI) relative to placebo.

[4] Indicators for assessing infant and young child feeding practices: definitions and measurement methods. Geneva: World Health Organization and the United Nations Children's Fund (UNICEF), 2021.
